# Navigating Life After COVID-19: Adoption of Preventive Behaviour Among Citizens Post-Pandemic

**DOI:** 10.1101/2025.04.29.25326653

**Authors:** Yifei Dong, Wong Chee Hoo, SPR Charles Ramendran, Pradeep Paraman

**Affiliations:** Universiti Malaya, 50603 Kuala Lumpur, Wilayah Persekutuan Kuala LumpurMalaysia; Inti International University, 71800 Nilai, Negeri Sembilan, Malaysia; Universiti Tunku Abdul Rahman, 31900 Kampar, Perak, Malaysia; Segi University, Research and Innovation Management Centre (RIMC), Pju 5 Kota Damansara, 47810 Petaling Jaya, Selangor,Malaysia

**Keywords:** Preventive Behaviours, COVID-19, Health Communication, Theory of Planned Behavior, Public Health Interventions, Mediating Role of Behavioural Intention

## Abstract

This study examines the determinants affecting the adoption of preventive behaviours among Malaysian individuals in the post-pandemic environment of COVID-19. The pandemic has profoundly affected public health, making the comprehension of these aspects essential for formulating effective health communication and intervention methods. This research aims to investigate how psychological and social factors, including attitudes (AT), subjective norms (SN), perceived behavioral control (PBC), perceived susceptibility (PSU), and perceived severity (PSE), forecast the adoption of preventive behaviors. A quantitative research approach was employed to survey a sample of 485 respondents in order to investigate the correlations between these parameters and the actual preventative behaviours. Data analysis was performed using structural equation modeling (SEM) to examine the direct and indirect correlations among the variables. The findings indicate that personal experiences and views, molded by the epidemic, substantially affect adherence to health guidelines. Furthermore, behavioral intention was identified as a mediator between the predictions and actual behaviors. This study addresses gaps in the literature and underlines the influence of digital platforms on public health behaviors. The results offer actionable recommendations for policymakers to improve the adoption of preventive behaviors and bolster public health responses, fostering a more resilient healthcare system in Malaysia.

## Introduction

The extraordinary severity of the coronavirus disease 2019 (COVID-19) presents a major public health issue. The post−COVID-19 condition (PCC) is a multifaceted and dangerous disorder that has impacted many lives worldwide. Recognizing the potential risk factors is crucial for understanding who may develop PCC, as it facilitates timely and effective treatment intervention [1]. Upon the emergence of COVID-19, the government swiftly launched a public health response and ensured sufficient medical care to address the public health crisis while enforcing movement restrictions nationwide beginning on 18 March 2020 [2].

In reaction to the extensive effects of COVID-19, the Malaysian government instituted multiple steps to protect public health [3, 4, 5]. However, the proliferation of fake news and disinformation during the pandemic underscores the need for effective risk communication strategies [6]. Addressing the pandemic requires a robust understanding of the factors influencing preventive healthcare behaviour. This behaviour, which encompasses actions taken to avert lifestyle diseases, is shaped by a range of personal, demographic, cultural, social, and socioeconomic factors. The successful adoption of these preventive measures is linked to positive outcomes including enhanced quality of life, consumer well-being, and effective health planning [7].

In Figure 1, the adoption of preventive behaviors is a central focus of this study, and understanding the historical context of how different Malaysian states managed the COVID-19 pandemic provides critical insights. The varying infection rates across states—such as Kedah in the northern region, Selangor and the Federal Territory (Kuala Lumpur) in the central region, Johor in the southern region of West Malaysia, and Sarawak in East Malaysia—reveal distinct challenges and responses to the pandemic. These differences raise important questions about how psychological and social factors, such as attitudes, perceived severity of the virus, and perceived behavioral control, were influenced by the states’ unique experiences during the crisis.

**Figure 1.:**
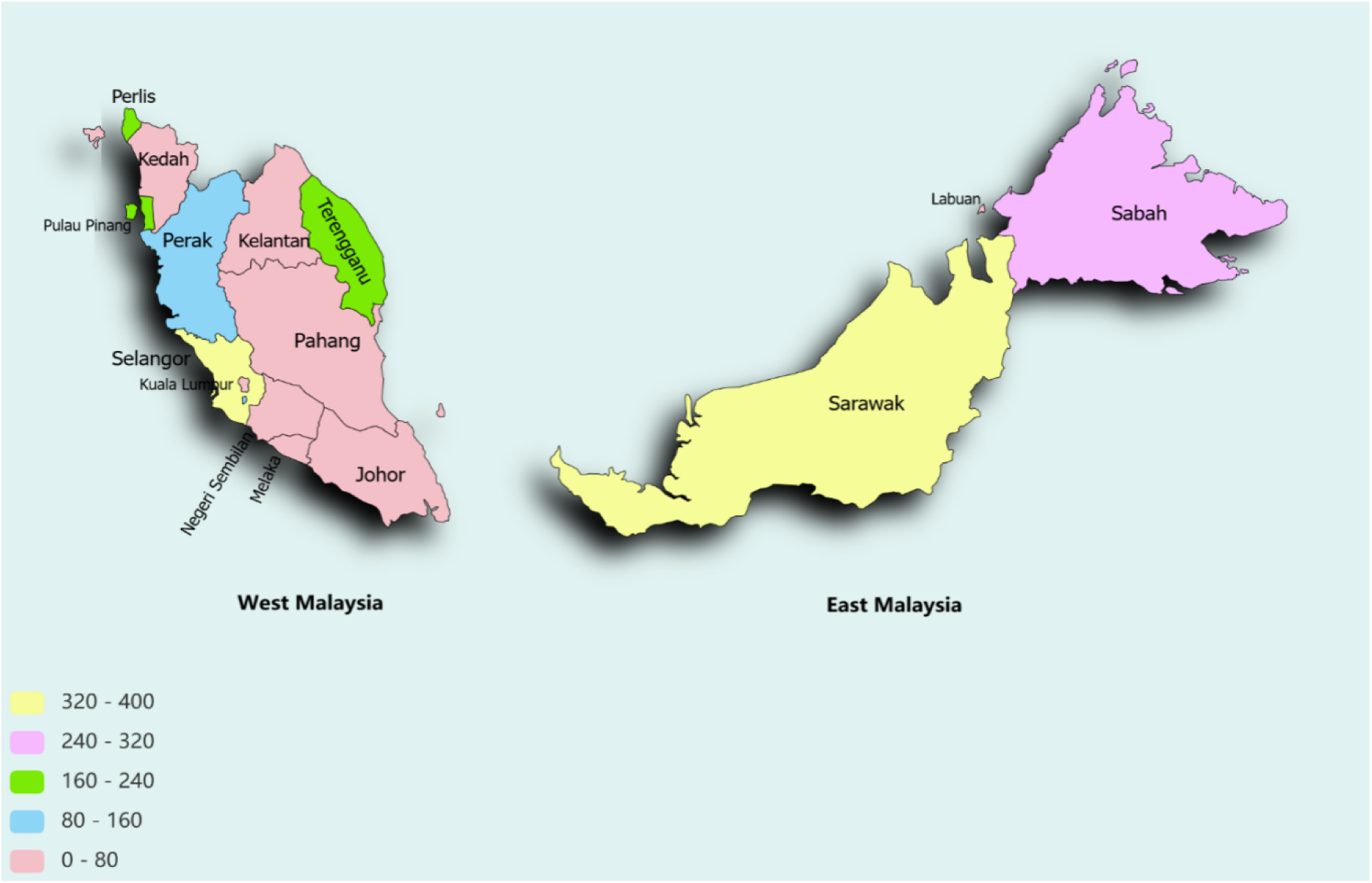
Regional COVID-19 Infection Rates Across Malaysian States [Source: Authors].

The table below (see Table 1) categorizes Malaysian states based on their COVID-19 infection rates, providing a clear overview of the pandemic’s impact across different regions. The infection rates are classified into five categories: Low (0-80), Moderate (80-160), High (160-240), Very High (240-320), and Critical (320-400). These categories reflect the intensity of COVID-19 transmission and highlight the varying levels of pandemic management effectiveness across states.

**Table 1:**
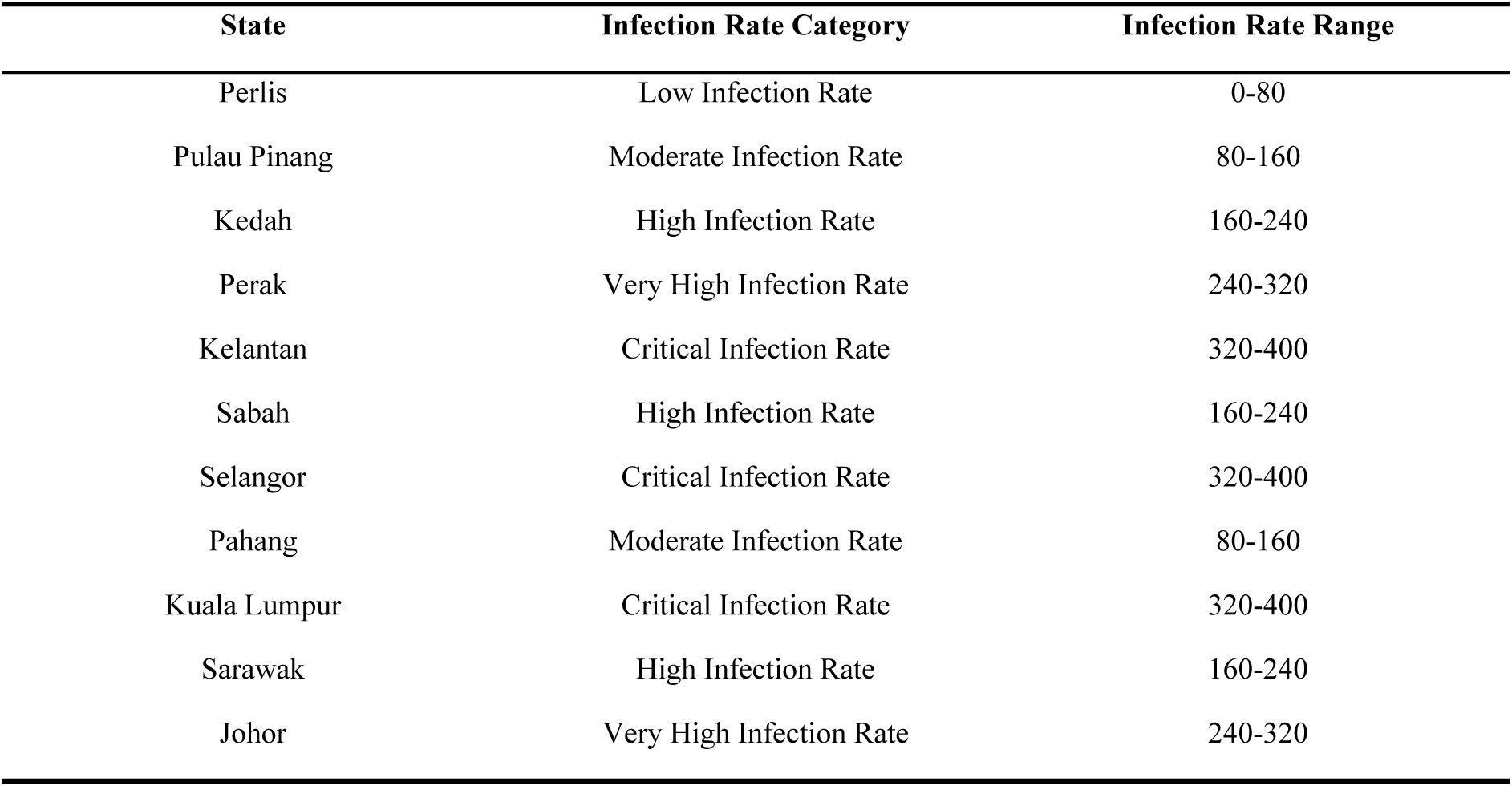
State Infection Rate Category and Range.

| State | Infection Rate Category | Infection Rate Range |
| --- | --- | --- |
| Perlis | Low Infection Rate | 0-80 |
| Pulau Pinang | Moderate Infection Rate | 80-160 |
| Kedah | High Infection Rate | 160-240 |
| Perak | Very High Infection Rate | 240-320 |
| Kelantan | Critical Infection Rate | 320-400 |
| Sabah | High Infection Rate | 160-240 |
| Selangor | Critical Infection Rate | 320-400 |
| Pahang | Moderate Infection Rate | 80-160 |
| Kuala Lumpur | Critical Infection Rate | 320-400 |
| Sarawak | High Infection Rate | 160-240 |
| Johor | Very High Infection Rate | 240-320 |

The COVID-19 infection rates across Malaysian states (see Table 1) varied significantly, reflecting diverse challenges and responses to the pandemic. These rates are categorized into five levels: Critical (320-400), Very High (240-320), High (160-240), Moderate (80-160), and Low (0-80). Each category provides insights into the intensity of transmission and the effectiveness of pandemic management efforts.

States with Critical Infection Rates (320-400), such as Selangor, Kuala Lumpur, and Kelantan, experienced the highest infection rates, indicating significant challenges in controlling the virus. These regions likely faced issues related to high population density, mobility, and varying levels of compliance with health guidelines. Similarly, states with Very High Infection Rates (240-320), including Perak and Johor, reflected substantial transmission, necessitating targeted interventions to curb the spread.

States categorized under High Infection Rates (160-240), such as Kedah, Sabah, and Sarawak, faced localized outbreaks, suggesting the need for focused public health efforts to address specific hotspots. In contrast, states with Moderate Infection Rates (80-160), like Pulau Pinang and Pahang, demonstrated relatively better control but still required vigilance to prevent further spread. Finally, Perlis, with its Low Infection Rate (0-80), highlighted successful containment efforts or lower transmission in the region.

The differing infection rates highlight the necessity of customized public health measures that take into account local settings, including population density, healthcare infrastructure, and socio-economic considerations. States with critical and very high infection rates may require intensified efforts, including enhanced testing, vaccination campaigns, and stricter enforcement of health guidelines. Meanwhile, regions with moderate to low infection rates should focus on sustaining their efforts to prevent future outbreaks, particularly through community engagement and effective risk communication.

By understanding these regional disparities, policymakers and public health officials can design targeted interventions to address the unique challenges faced by each state, ultimately improving Malaysia’s resilience to future public health crises.

Despite previous studies identifying the direct effects of AT and SN on the intention to adopt preventive behavior, gaps remain in comprehensively exploring the roles of PBC, PSU, and PSE in the Malaysian context. In addition, the influence of digital platforms—such as e-government information and social media—on shaping attitudes and intentions has not been thoroughly investigated. Moreover, the mediating effect of behavioural intention among these factors is underexplored, highlighting the need for a nuanced understanding of how these variables interact to impact preventive behaviour adoption.

Thus, this study focuses on three research questions:

1. What is the level of adoption of COVID-19 preventive behaviour among Malaysian citizens post-pandemic?
2. How do AT, SN, PBC, PSU, and PSE influence the intention to adopt preventive behaviour among Malaysian citizens?
3. Does behavioural intention mediate the relationships between these factors and the actual adoption of preventive behaviour?

By addressing these questions, the study aims to explore the adoption of COVID-19 preventive behaviour among Malaysian citizens. The first question provides a foundational understanding of public compliance with health guidelines. The second delves into the psychological and social factors influencing individuals’ intentions, examining the interactions of these determinants. Finally, the third question investigates the mediating role of behavioural intention, clarifying how intentions translate into actions.

### Problem Statement

The COVID-19 pandemic highlighted the vital role of preventive behaviors in curbing public health crises. However, as societies transition into post-pandemic phases, critical gaps persist in understanding how these behaviors are sustained, particularly in culturally diverse and digitally engaged nations like Malaysia.

First, while extensive research has explored factors influencing pandemic-era compliance—such as knowledge[3], attitudes[4], and digital tools[8–13]—most studies adopt a fragmented approach. They examine these factors in isolation, neglecting how health system performance, psychological drivers, and social norms interact to shape long-term behavior. For instance, Hamzah et al. [2] assessed Malaysia’s health system efficiency but did not investigate how public trust in the system combines with perceived risk to sustain preventive actions. This study integrates these elements into a unified framework grounded in the Theory of Planned Behavior (TPB) and Theory of Reasoned Action (TRA).

Second, although behavioral intention is central to TPB/TRA, its role as a mediator between external predictors (e.g., policy changes, digital nudges) and actual behavior remains under-tested, especially in post-crisis contexts. In Malaysia, where centralized health messaging and collective social values may amplify or weaken this mediation, understanding this linkage is essential to design effective interventions.

Third, prior work largely overlooks contextual nuances specific to Malaysia, such as urban-rural disparities in healthcare access, ethnic diversity in risk perception, and the long-term use of digital platforms like MySejahtera. These gaps limit the applicability of findings to real-world policy design.

This study bridges these gaps by:

1. Proposing the first integrated model that connects health system performance, digital engagement, and psychosocial factors to explain post-pandemic behaviour.
2. Empirically testing how behavioural intention mediates the effects of external drivers (e.g., trust in government alerts) on sustained compliance.
3. Providing locally actionable insights for Malaysia, with implications for other multi-ethnic, digitally transitioning societies.

By addressing these challenges, the research advances theoretical understanding of resilient health behaviours and equips policymakers to tailor strategies for future public health threats.

### Literature Review

This literature analysis examines the effectiveness of risk communication and collaborative partnerships in addressing the pandemic and highlights areas lacking current research. Through analyzing various communication tactics and collaborations, we aim to discover valuable lessons that can guide future public health strategies. Moreover, this study emphasizes the need for additional, comprehensive research on the post-pandemic phase, particularly in understanding the changes in behavior and enhancing communication strategies for future emergencies.

It is, therefore, essential to use frameworks like the Theory of Planned Behavior (TPB) and the Health Belief Model (HBM). HBM describes how people perceive risks, mainly severity and vulnerability, and perception towards health-related behaviors. Essentially, this is the framework concerning the effect that risk communication may have on the behavior of the public during this pandemic on compliance with existing health guidelines. TPB informs how behavioral intentions are formed and their consequences for actual behaviors. The literature review will call on this framework to understand the protocols used during risk communication and their effect on behavioral changes during and after the pandemic.

The literature review discusses the Malaysian government’s shortcomings in its risk communication strategy during the first COVID-19 phase and the government’s solution to correct its shortcomings. This section discusses the gaps that came up through the first communication strategy the government used to communicate COVID-19 and how effective MCO was in solving the identified gaps. It examines the effectiveness of the Malaysian government in implementing MCO and how it may have changed the citizen’s behavior. This section gives valuable insights that governments can use for effective risk communication during pandemics.

#### Overview of Risk Communication in Malaysia

During and after the COVID-19 pandemic, Malaysia faced significant challenges as a level three warning nation under the Centers for Disease Control and Prevention [14]. Preventive measures included social distancing, lockdowns, and educational postponements, but the government encountered difficulties in identifying and isolating cases. Effective risk communication, led by the Ministry of Health and the National Security Council, was instrumental. This involved press briefings, social media outreach, and public service announcements, often in collaboration with community leaders and health professionals to ensure culturally relevant communication [15].

Efforts to engage remote communities highlighted the importance of tailored strategies to overcome communication barriers [15]. Research from Bangladesh similarly emphasized challenges, noting disparities in access to information between urban and rural women. Rural women had better support networks despite equal challenges, while urban low-income women struggled due to limited technology access and communication with authorities [16].

Lessons from Malaysia suggest that transparent communication, collaboration with medical experts, and engagement with community leaders were critical in managing the crisis [17]. A comprehensive communication strategy, with consistent messages aligned with Malaysia’s cultural mission, reduces misunderstandings and enhances public satisfaction [18]. To improve future crisis communication, Malaysia should enhance its communication infrastructure, ensure timely and accurate information dissemination, involve stakeholders across sectors, and actively address misinformation.

#### Successful Risk Communication Strategies

Malaysia’s Stay Safe, Home Campaign proved highly effective in promoting public compliance with lockdown measures. By delivering consistent messages through social media, television, and radio, the government ensured widespread understanding. The use of videos and infographics further facilitated comprehension, making it easier to reach broader audiences.

Daily press briefings by the Director-General of Health offered timely and transparent updates, instilling public confidence [19]. These sessions provided comprehensive information on new cases, recovery rates, and ongoing initiatives. The government’s strategic use of social media platforms like Facebook and Twitter helped swiftly counter misinformation, share health guidelines, and directly engage the community [19].

#### Less Successful Risk Communication Strategies

Contradictory messaging around vaccine benefits and potential side effects caused public confusion and hesitancy [19]. Early communication failed to adequately address concerns about vaccine safety, undermining public trust. Language barriers also presented challenges. Communities that did not speak Malay often lacked access to translated and culturally relevant health messages. This gap hindered the effective dissemination of information.

Additionally, insufficient engagement with rural areas further complicated communication. Rural communities with limited internet connectivity were often neglected, resulting in less awareness of health guidelines and lower adherence to protocols [20].

### Literature Gaps

The literature review reveals that while previous research has explored crisis communication during the pandemic, effective communication strategies, and approaches to curb COVID-19, there are significant gaps in the literature regarding the relationship between risk communication campaigns and citizen behavior, especially in Malaysia. This study aims to fill these gaps by focusing on the effectiveness of government risk communication and its impact on the adoption of preventive behavior after the COVID-19 pandemic.

In a systematic review of the literature encompassing 82 studies worldwide on the predictors of preventive behaviours related to COVID-19 [21], only two studies were conducted in Malaysia: Thong et al. [23] and Mat Dawi et al. [3]. Thong et al. [23] applied the HBM to examine how perceptions of benefits, susceptibility, and barriers influenced adults’ intentions to receive the vaccine. Mat Dawi et al. [4] utilized the TRA to explore the relationship between attitudes and subjective norms in shaping intentions towards preventive behaviours.

Past research, such as Rowan et al.’s use of the Health Belief Model (HBM) for cancer risk communication (2003) and training officials in risk communication (2009), primarily focused on officials’ perspectives. However, VanDyke and King (2018) suggested that future research should examine public perspectives and international contexts [24]. This study applies the Health Belief Model in Malaysia, broadening its scope beyond the U.S.

Additionally, the TPB developed by Ajzen has been widely used to explore behavioral intentions, often incorporating additional factors [25]. This study combines TPB with HBM to create a more comprehensive framework for understanding behavior during crises. The integration of TPB’s key factors (AT, SN, and PBC) with HBM’s focus on health beliefs is expected to provide deeper insights into the behavioral factors that influence preventive actions during pandemics.

This holistic approach emphasizes the importance of addressing perceived barriers, highlighting the benefits of preventive measures, and understanding the social consequences of public health communication. Policymakers and health authorities can use this framework to refine communication campaigns, enhance compliance with health guidelines, and ultimately improve crisis management strategies.

Furthermore, this study identifies key gaps in the literature, particularly the lack of post-pandemic studies and the need for deeper investigation into how socio-economic factors, education, and cultural backgrounds affect responses to risk communication. Additionally, there is limited research on the effectiveness of hybrid communication strategies that combine traditional and digital media. This study aims to fill these gaps and provide valuable insights for managing future public health crises.

### Theoretical Rationale

Behavioral change theories and models comprise a collection of interconnected concepts, definitions, and propositions that define, explain, predict, or regulate behaviors [26]. They are employed to comprehend the reasons individuals engage in or abstain from health-promoting behaviors, ascertain the requisite information for formulating successful intervention techniques, and establish priorities for health education and promotion initiatives [27]. The HBM, TRA (Theory of Reasoned Action), and TPB, transtheoretical model, extended parallel process model, socio-ecological model, social cognitive theory, and diffusion of innovations theory are the most frequently employed in behavioral change intervention tactics. HBM is employed to comprehend the reluctance of individuals to embrace disease preventative measures or undergo screening procedures for early illness diagnosis. The model has six constructs: perceived vulnerability, perceived severity, perceived barrier, perceived benefit, self-efficacy, and cues to action [28]. The TPB and the TRA assert that behavioral outcomes are contingent upon behavioral intentions. The components of the TRA (attitude and subjective norms) predicted COVID-19 preventative behaviors, including handwashing, mask usage, and COVID-19 vaccination [29]. The Transtheoretical Model (TTM) emphasizes individual decision-making for purposeful change, encompassing five stages: pre-contemplation, contemplation, preparation, action, and maintenance.

Our study’s focus on the TPB and HBM is well justified for several reasons. First, both are established frameworks that directly address the relationship between beliefs, intentions, and behaviours. Given our aim to investigate the predictors of preventive behaviours, these theories provide a solid foundation for understanding how attitudes and subjective norms influence intentions, which affect actual behaviour, particularly in the context of health behaviours related to COVID-19.

Moreover, while the TPB includes attitudes, subjective norms and perceived behavioural control as a significant predictor, the HBM focuses on perceived susceptibility and perceived severity, thereby enabling our study to capture a broad range of psychological and social factors influencing behavioural intentions. This is particularly relevant in addressing gaps in the existing literature regarding the roles of perceived susceptibility and perceived severity, which are critical for understanding health-related behaviours. Integrating these constructs into the analysis enriches understanding of how citizens perceive their risk and implications of their behaviour.

In addition, the focus on behavioural intention as a mediator in the relationship between various predictors and actual behaviour is crucial for public health interventions. This study contributes new insights into hitherto underexplored areas of research, enhancing the theoretical understanding of behaviour change. The selection of the TPB and HBM is also suitable given their adaptability across different cultural contexts, making them relevant in the Malaysian setting, where cultural and social factors significantly influence health behaviours.

Ultimately, employing the TPB and HBM advances theoretical knowledge and provides actionable insights for policymakers. By understanding the interplay between attitudes, subjective norms, and behavioural control, our study can inform effective health communication strategies tailored to the Malaysian populace, especially in terms of combating misinformation and promoting health compliance. In summary, applying the TPB and HBM enables addressing the multifaceted nature of health behaviours to offer a comprehensive framework, (see fig.2), that captures the complexities of behaviour change in a post-pandemic context while filling existing research gaps. The hypotheses (see table 2 and 3) are presented below.

**Figure 2:**
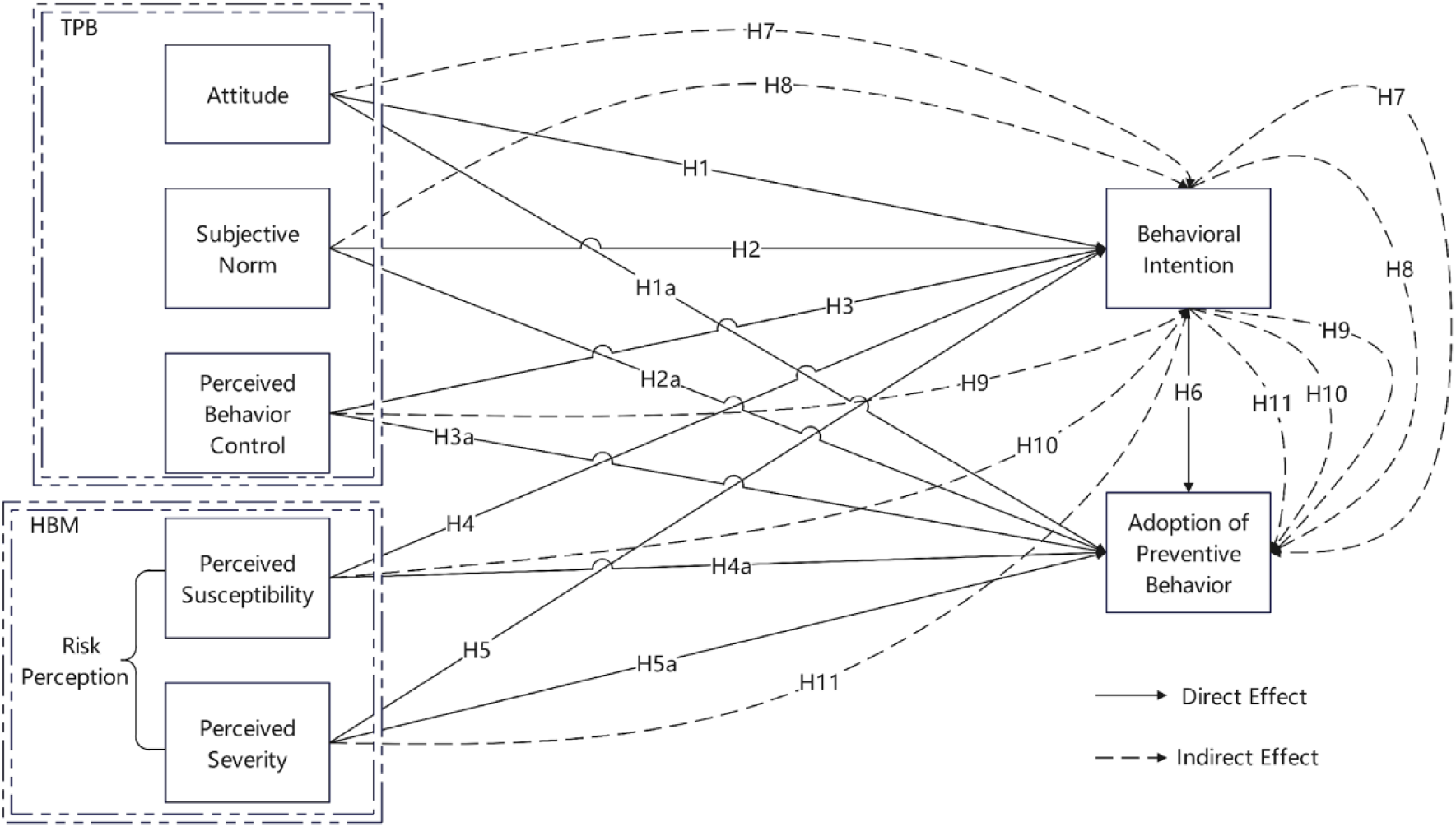
Conceptual Framework.

**Table 2:**
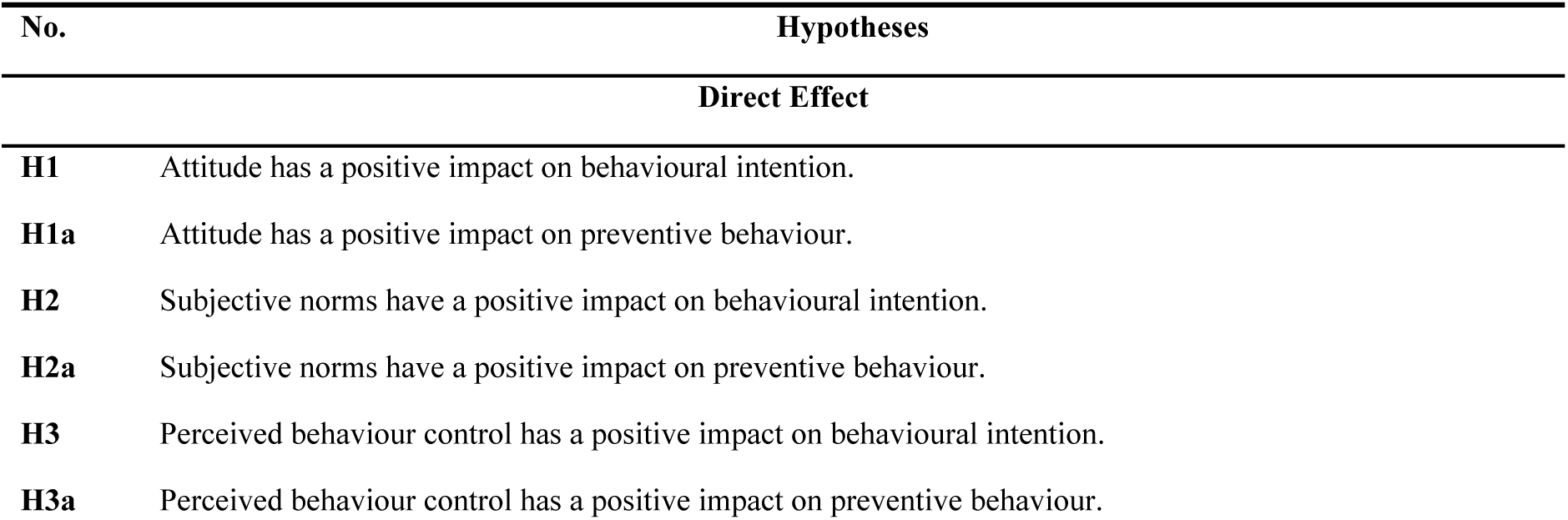
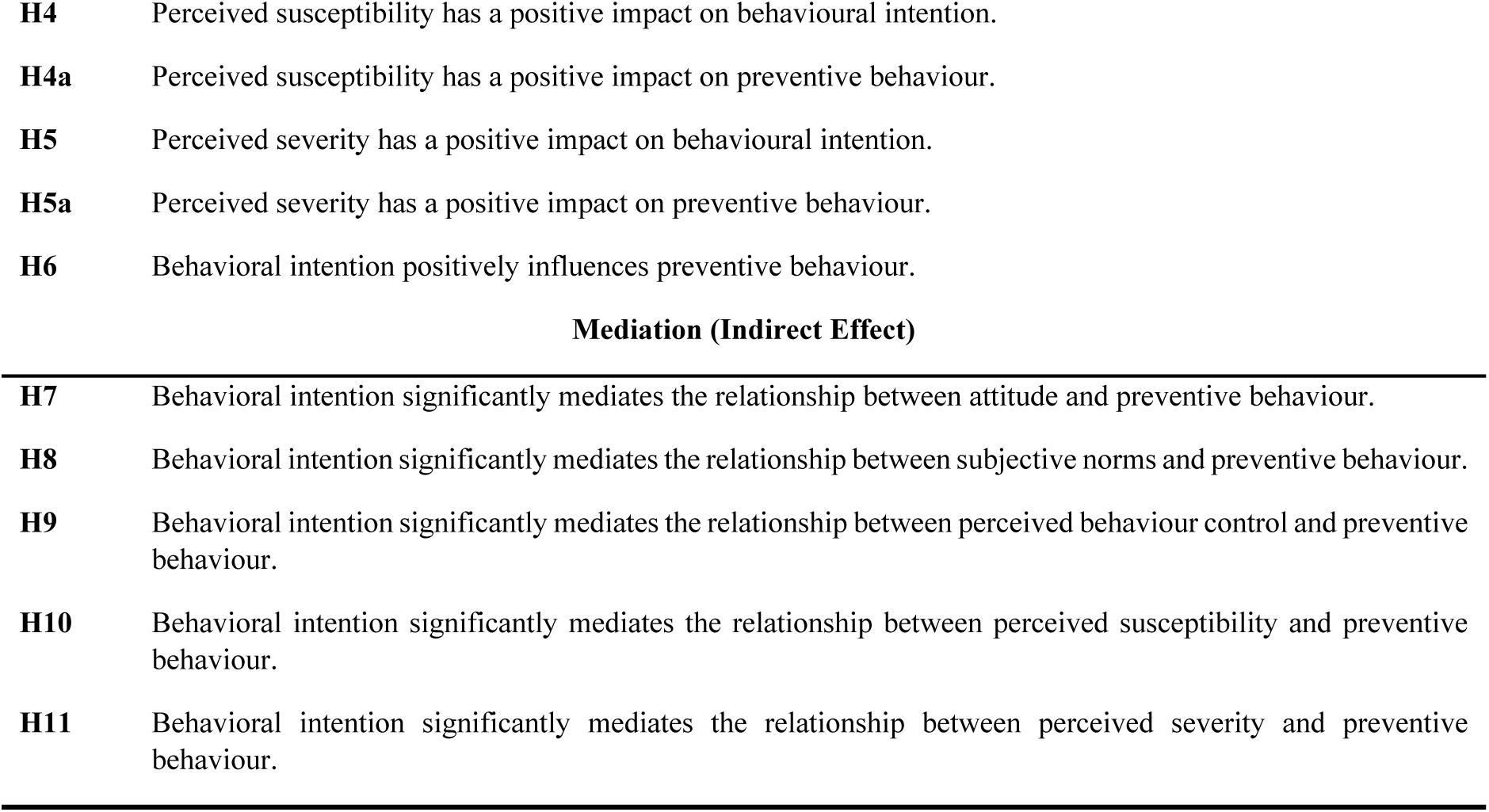
Hypotheses - Direct and Indirect Effect.

**Table 3:** Hypotheses – Effects and Theoretical Underpinning.

| Hypotheses | Statement | Theoretical Support |
| --- | --- | --- |
|  | Direct Effect | Theoretical Underpinning |
| H1 | Attitude has a positive impact on behavioural intention. | Supported by the TPB [25], which posits that attitudes toward a behavior strongly influence individuals' intentions to engage in that behavior. |
| H1a | Attitude has a positive impact on preventive behaviour. | Empirical studies suggest that individuals with positive attitudes towards preventive health behaviors are more likely to adopt such behaviors [27]. |
| H2 | Subjective norms have a positive impact on behavioural intention. | Social influences play a critical role in shaping behavioral intentions, as highlighted by the TPB [29]. |
| H2a | Subjective norms have a positive impact on preventive behaviour. | Perceptions of social pressure can drive individuals to adopt preventive behaviors [30]. |
| H3 | Perceived behavioral control has a positive impact on behavioural intention. | Greater perceived control over a behavior increases the likelihood of intention to perform that behavior [25]. |
| H3a | Perceived behavioral control has a positive impact on preventive behaviour. | When individuals feel empowered to engage in preventive behaviors, they are more likely to do so [31]. |
| H4 | Perceived susceptibility has a positive impact on behavioural intention. | The Health Belief Model [32] posits that persons who recognize an elevated risk of health problems are more inclined to develop preventative intentions. |
| H4a | Perceived susceptibility has a positive impact on preventive behaviour. | High perceived vulnerability often leads to the adoption of health-protective behaviors [33]. |
| H5 | Perceived severity has a positive impact on behavioural intention. | When individuals believe that a health issue is severe, they are more likely to develop preventive intentions [32]. |
| H5a | Perceived severity has a positive impact on preventive behaviour. | The perception of severe consequences can motivate preventive action [33]. |
| H6 | Behavioral intention positively influences preventive behaviour. | Strong intentions have been consistently linked to actual behavioral execution [25]. |

|  | Indirect Effect (Mediation) | Theoretical Causality Paths |
| --- | --- | --- |
| H7 | Behavioral intention significantly mediates the relationship between attitude and preventive behaviour. | Intentions serve as the bridge between attitudes and actual behaviors [25]. |
| H8 | Behavioral intention significantly mediates the relationship between subjective norms and preventive behaviour. | Social norms influence behavior primarily through the formation of intentions [25]. |
| H9 | Behavioral intention significantly mediates the relationship between perceived behavioral control and preventive behaviour. | Perceived control impacts behavior indirectly through intention [25]. |
| H10 | Behavioral intention significantly mediates the relationship between perceived susceptibility and preventive behaviour. | Perceived risk affects behavior through the development of preventive intentions [33]. |
| H11 | Behavioral intention significantly mediates the relationship between perceived severity and preventive behaviour. | The belief in severe health outcomes shapes behavior via intention formation [32]. |

Figure 2 illustrates the framework that amalgamates TPB [25] and HBM [34] to examine the behaviors of Malaysian residents. Alagili and Bamashmous [34] have shown that HBM effectively elucidates COVID-19 preventative strategies, whereas the TPB has been extensively utilized to examine behavioral reactions in many crisis scenarios. The framework (refer to Fig. 2) underscores the importance of the HBM and the TPB in comprehending and facilitating effective communication. The HBM asserts that individuals’ behavioral intentions are profoundly affected by their perceptions of vulnerability, severity, benefits, and obstacles. These objectives subsequently inspire specific preventive measures. The TPB elucidates this understanding by incorporating AT, SN, and PBC as essential determinants of behavioral intentions. Effective communication must meticulously evaluate and handle these issues to influence citizens’ perspectives and intents. The approach emphasizes the necessity of well-planned risk communication strategies to foster public compliance with health regulations, hence improving overall public health outcomes during pandemics like COVID-19 [35]. This study’s originality stems from its thorough and integrated examination of the adoption of preventative behaviors among Malaysian residents in a post-pandemic setting. This research broadens the focus of previous studies that examined particular constructs within established behavioral models by integrating multiple predictors—attitudes, subjective norms, perceived behavioral control, perceived susceptibility, and perceived severity—into a cohesive framework. This method is for a more detailed examination of how various elements interact and jointly affect behavioral intentions.

Moreover, the study addresses significant gaps in the existing literature, particularly regarding the role of perceived susceptibility and perceived severity, which have been less explored in the Malaysian context. By investigating the mediating effect of behavioural intention, this research contributes a deeper understanding of how intentions translate into actual preventive behaviours. It also examines the influence of digital platforms such as e-government and social media on shaping health behaviours, an aspect largely overlooked in prior studies.

The limited representation of Malaysian research in the global context of pandemic preventive behaviour studies highlights the need for more localized investigations. This paper not only fills this gap, but also provides actionable insights for public health interventions, making a significant contribution to the understanding of health behaviour adoption in Malaysia and beyond.

### Methodology

This section delineates the research technique employed to examine preventative health behaviors among Malaysian residents in the post-COVID-19 setting. The research used a quantitative methodology, incorporating the HBM and the TPB to analyze the correlations among principal variables. Data was gathered using a standardized questionnaire, with pre-testing and pilot testing performed to guarantee reliability and validity. Structural Equation Modeling of PLS (PLS-SEM) was employed for analysis due to its appropriateness for intricate models involving latent constructs. The chapter delineates the research design, sample methods, data collection procedures, and analytical approaches to guarantee transparency and replicability.

#### Research Design

The research employed a quantitative design, emphasizing the collecting of primary data to yield context-specific insights. A structured questionnaire was used to systematically measure and numerically analyze variables. Descriptive analysis was employed to assess the current status of variables, and hypotheses were formulated after preliminary data analysis to ensure a data-driven approach. The methodology prioritized objectivity, rigor, and reliability, with results derived from statistical computations.

#### Sampling Procedure

Recruitment leveraged digital platforms, including social media and online advertisements, to maximize outreach. Screening questions verified eligibility, ensuring respondents met inclusion criteria. The sampling strategy combined inclusivity and precision, enabling a comprehensive analysis of preventive health behaviours while maintaining generalizability to similar urban contexts in Malaysia.

#### Instrument Development

The primary data collection instrument was a structured questionnaire, chosen for its efficiency and alignment with the study’s quantitative approach. The questionnaire had items utilizing a 5-point Likert scale to assess attitudes, perceptions, and preferences. It was divided into four main sections (see Table 4):

**Table 4:**
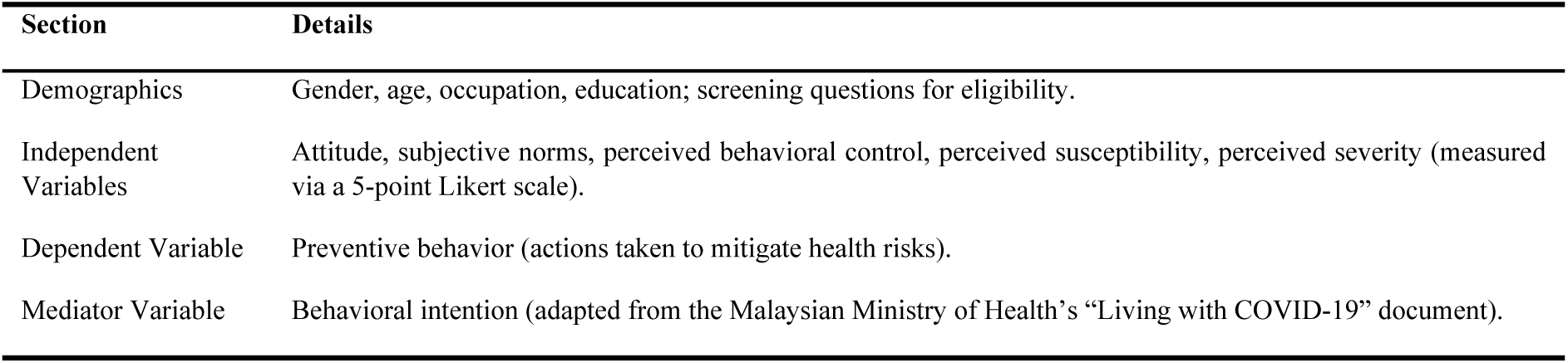
Questionnaire Structure.

#### Study Population and Unit of Analysis

The study population consists of Malaysian citizens aged 18 and above residing in Kuala Lumpur. This demographic is particularly relevant given the Malaysian Ministry of Health’s implementation of the “Living with COVID-19” guidelines, which outline preventive health behaviors for citizens.

The unit of analysis is the individual citizen, as risk perception and preventive behaviors are highly personal and subjective. Each citizen’s unique experiences and perceptions are critical to understanding the relationships between risk perception, behavioral intention, and preventive behavior.

#### Sample Size Determination

To determine the appropriate sample size, G*Power statistical software was used. With 11 predictors, 0.15 effect size, and a confidence level of 95%, the analysis indicated a minimum requirement of 178 respondents. To account for potential non-responses and enhance reliability, over 500 questionnaires were distributed, resulting in 485 valid responses. This exceeds the recommended sample size, ensuring robust and generalizable findings.

### Data Collection

A cross-sectional survey was performed utilizing a standardized, closed-ended questionnaire of 34 items. The questionnaire comprised seven components, each assessed using a 5-point Likert scale according to the framework (see Table 5).

**Table 5:**
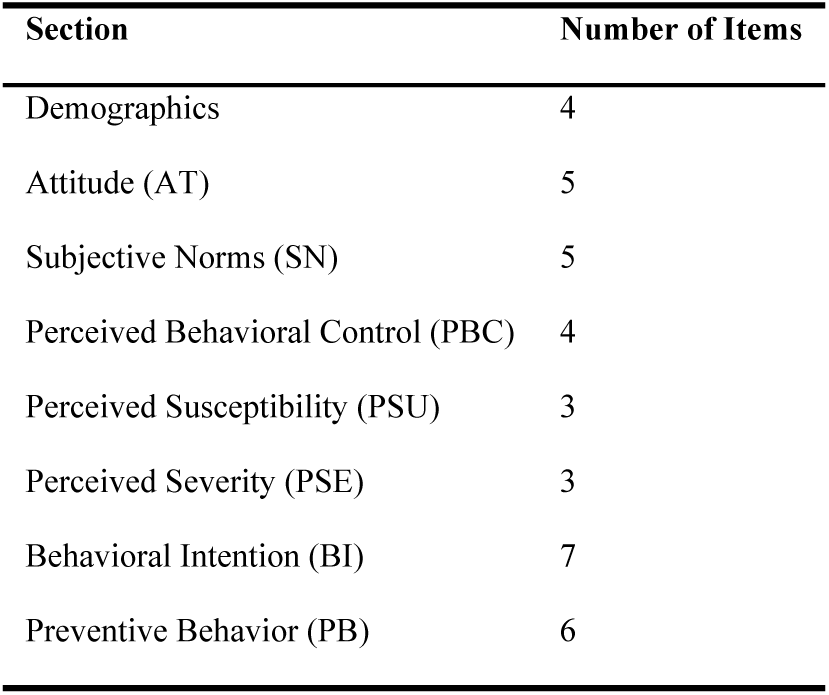
Questionnaire Structure.

| Section | Number of Items |
| --- | --- |
| Demographics | 4 |
| Attitude (AT) | 5 |
| Subjective Norms (SN) | 5 |
| Perceived Behavioral Control (PBC) | 4 |
| Perceived Susceptibility (PSU) | 3 |
| Perceived Severity (PSE) | 3 |
| Behavioral Intention (BI) | 7 |
| Preventive Behavior (PB) | 6 |

The questionnaire was administered online via Google Forms, with targeted advertisements on Google Ads and Meta Ads to reach the Kuala Lumpur population. Screening questions ensured participants met the inclusion criteria: aged 18 or older, residing in Kuala Lumpur, and Malaysian citizens. Responses failing these criteria were automatically excluded.

#### Pre-Test and Pilot Test

A pre-test (3–8 March 2024) identified ambiguities in the questionnaire using feedback from three expert reviewers. A pilot test (9–15 March 2024) with 30 participants assessed feasibility and refined the instrument.

#### Reliability and Validity

Reliability was evaluated by Cronbach’s alpha to determine internal consistency. A value of 0.7 or greater was deemed acceptable. The constructs assessed in the questionnaire—attitude, subjective norms, perceived behavioral control, perceived susceptibility, perceived severity, behavioral intention, and preventive behavior—exhibited high internal consistency, confirming the stability and reliability of the data (see table 5).

#### Validity Analysis During Pre-Tes*t*

Validity was ensured through multiple approaches to confirm that the questionnaire accurately measured the intended constructs. Content validity was achieved by aligning the questionnaire items with established studies [63, 64] and incorporating feedback from expert reviewers.

Construct validity was confirmed through factor analysis, which verified that the items accurately measured the theoretical constructs, such as attitude, subjective norms, perceived behavioural control, perceived susceptibility, perceived severity, behavioural intention, and preventive behaviour. Together, these measures ensured the questionnaire’s validity and its ability to generate meaningful and accurate data.

### Data Analysis

PLS-SEM was employed to evaluate the data due to its appropriateness for intricate models involving latent components. The analysis assessed hypothesized relationships between variables, providing robust insights into preventive health behaviours. Statistical computations ensured unbiased and reliable results, supporting the study’s objectives.

#### Demographic Profile of Respondents

The study collected data from 485 respondents, with the following demographic breakdown (see Figure 3). The figure provides descriptive statistics for the demographic variables of the study participants. The majority of participants were male (52.2%), while 47.8% were female. The most common age group was 25-34, representing 52.2% of the participants. The youngest age group was 18-24 (12.8%), and the oldest was 60 and above (1.4%). The most common level of education was high school/vocational school/technical school (39.4%), followed by college studies (18.4%). Only a small percentage of participants had a Ph.D. degree or above (1.4%). The most common occupation was employed (56.1%), followed by self-employed (26.2%). A relatively small percentage of participants were unemployed (14.2%).

**Figure 3:**
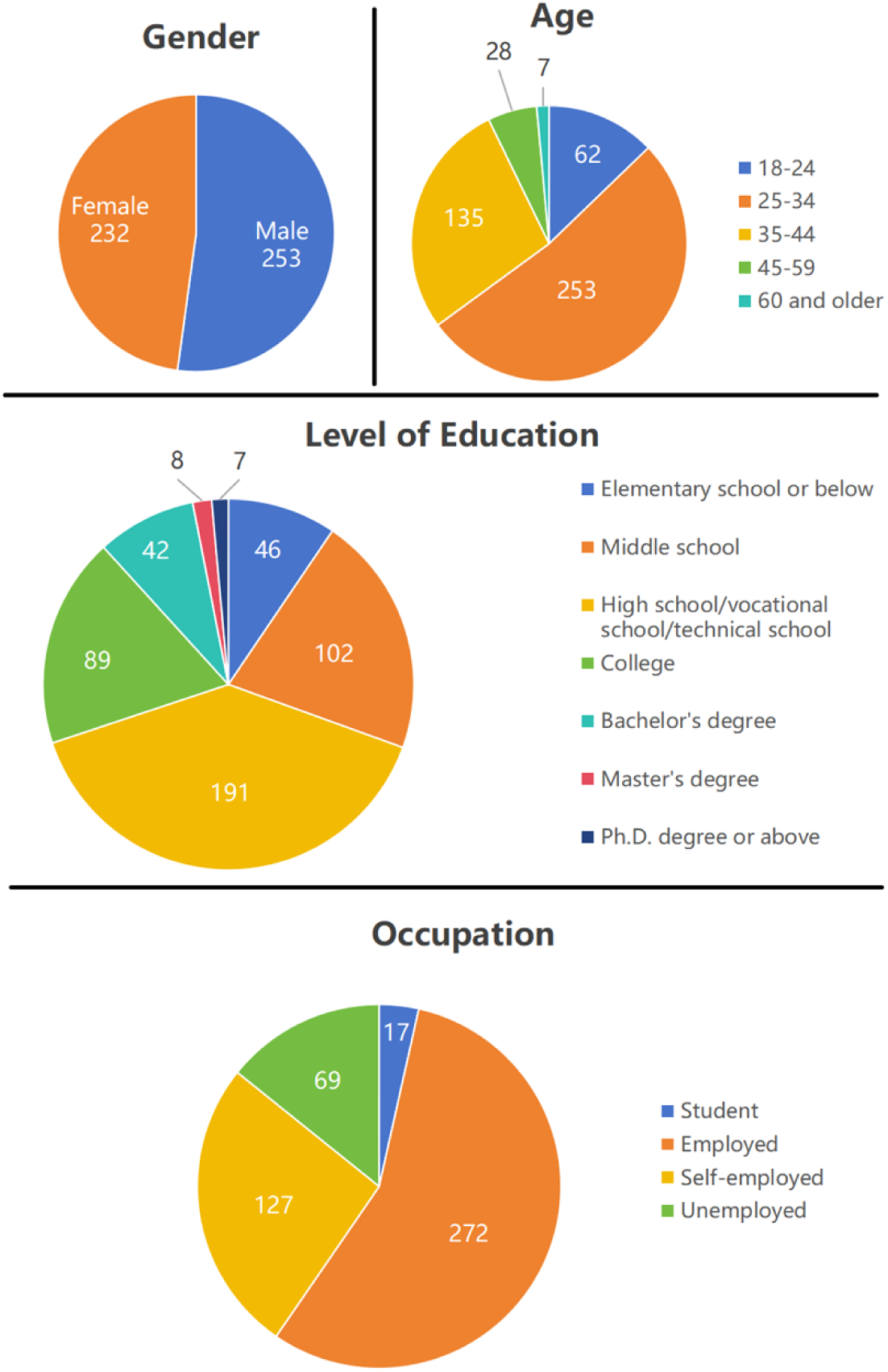
Demographic Information of Respondents.

#### Assessment and Goodness of Measurement Model

The measurement model in SmartPLS-4 serves as a statistical method for testing the validity and reliability of constructs or latent variables within SEM [36]. It tests the relationships between individual items and the construct that they refer to as well as among themselves. Figure 4 below illustrates the measurement model used in the present study.

**Figure 4:**
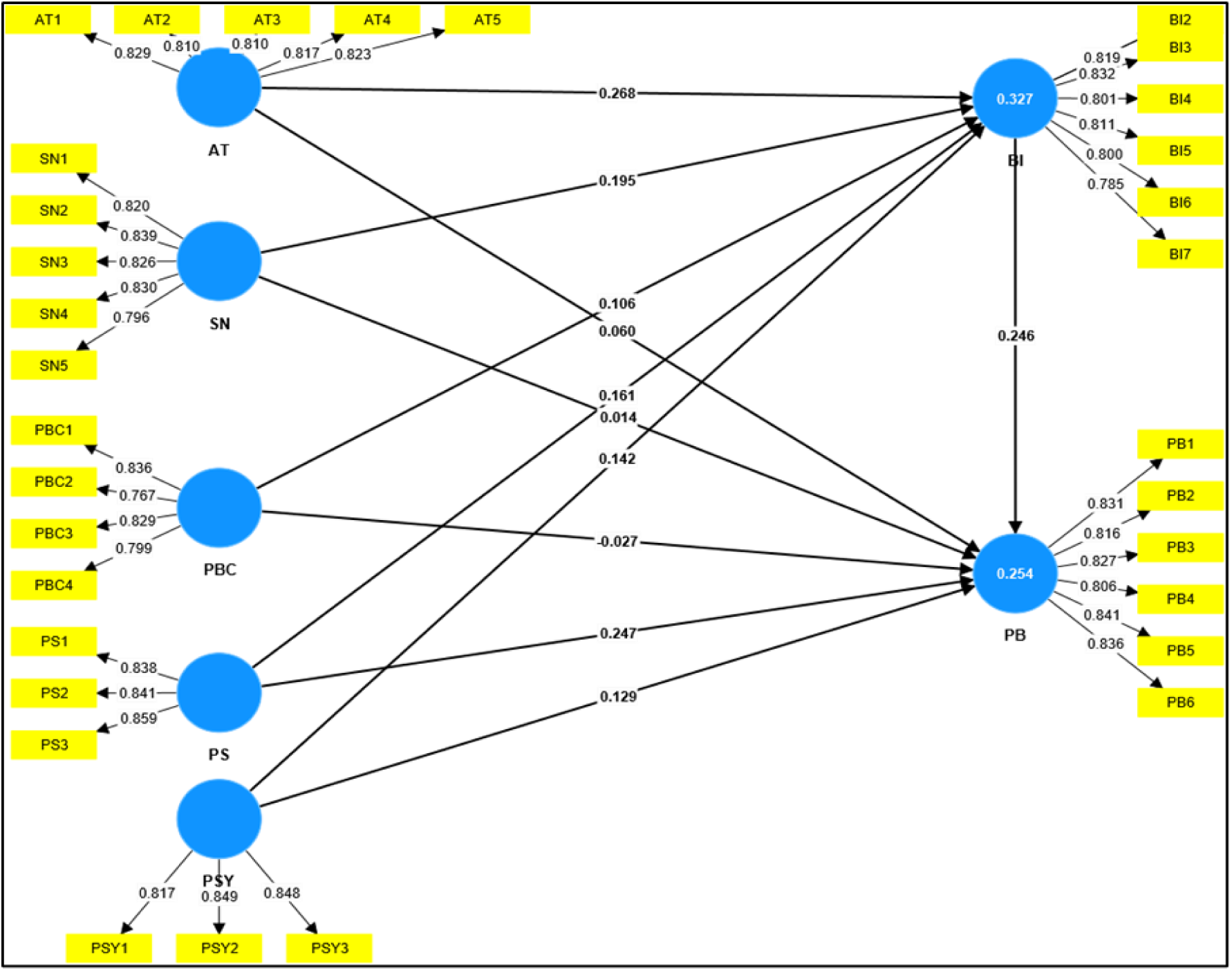
Measurement Model.

#### Measurement Model Assessment

The measuring model (refer to Figure 4) was assessed for convergent validity, reliability, and discriminant validity to confirm the robustness of the constructs.

Table 6 and Table 7 provide the convergent validity, including AVE values for each construct in the study. As demonstrated in the Table 7, the AVE values for all constructs exceed 0.50, confirming the convergent validity of the measurement model. This indicates that the constructs effectively capture the underlying phenomena of interest and that the indicators used to measure them are reliable and valid.

**Table 6:**
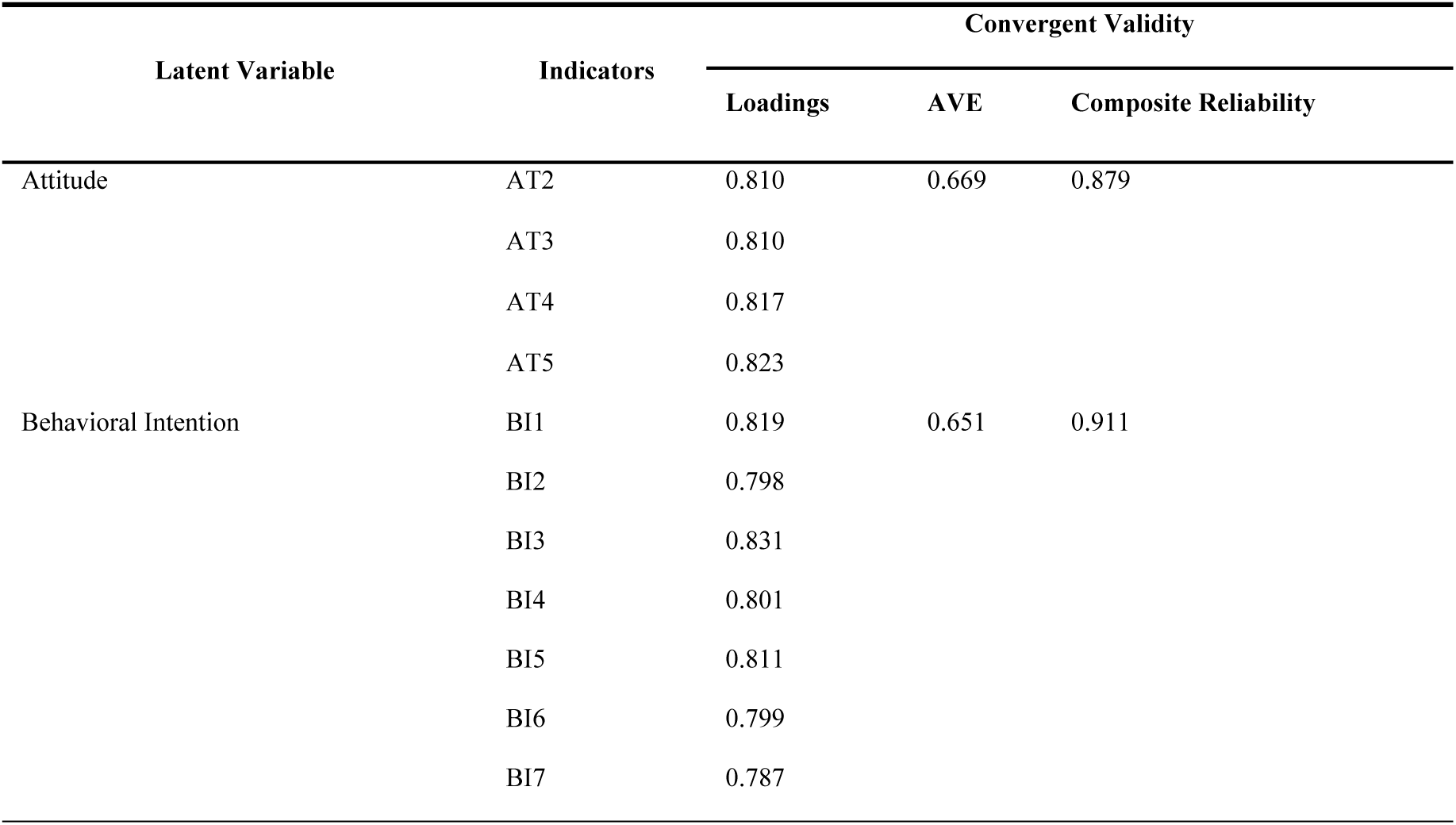
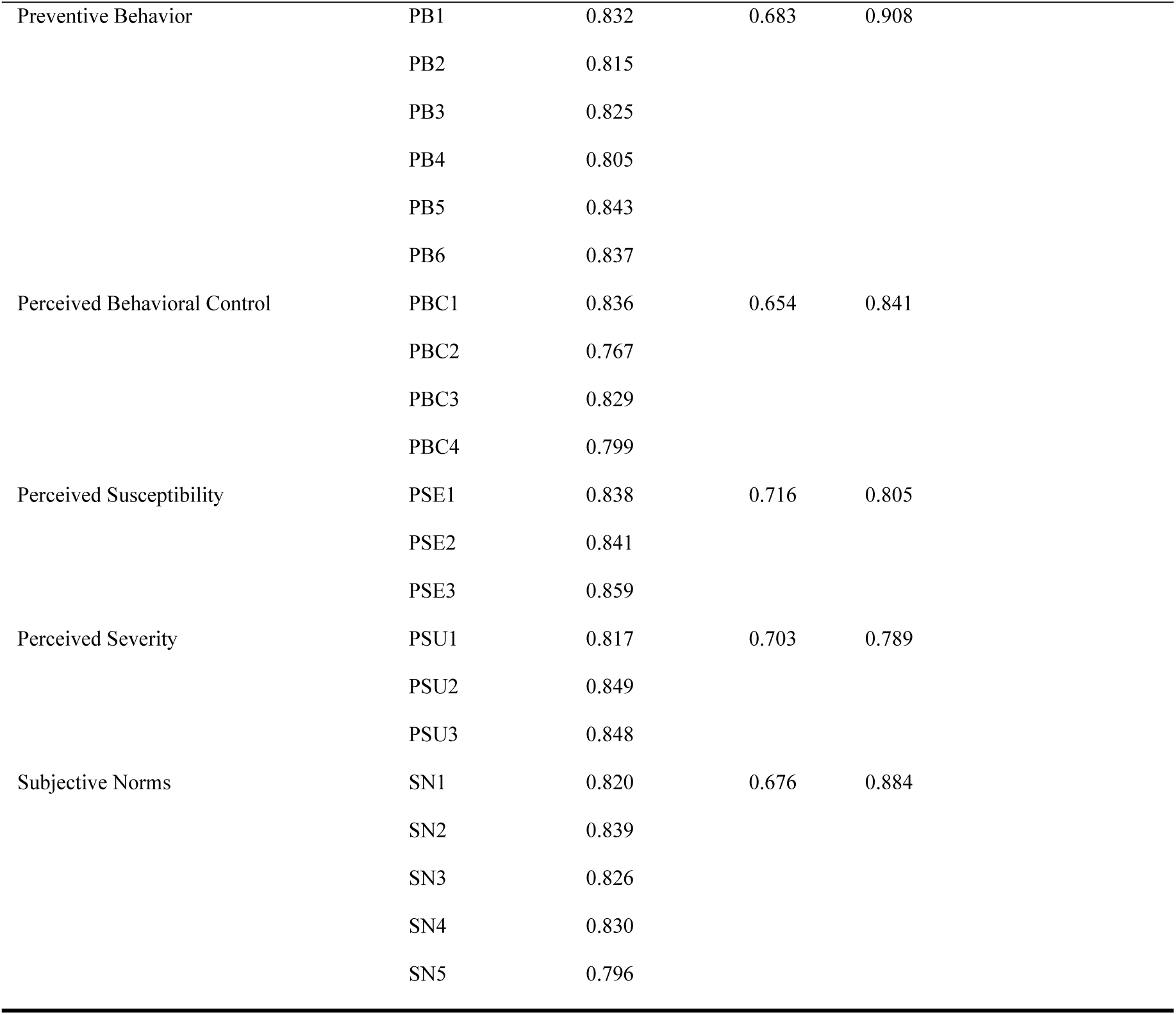
Measurement Model.

**Table 7:** Average Variance Extracted (AVE)

|  | Average variance extracted (AVE) |
| --- | --- |
| AT | 0.669 |
| BI | 0.651 |
| PB | 0.683 |
| PBC | 0.654 |
| PSE | 0.716 |
| PSU | 0.703 |
| SN | 0.676 |

#### Discriminant Validity

Discriminant validity guarantees that constructs are differentiated from each other. The evaluation was conducted utilizing the Fornell-Larcker Criterion and the Heterotrait-Monotrait (HTMT) Ratio. Table 8 delineates the Fornell-Larcker criteria.

**Table 8:** Fornell-Larcker Criterion.

|  | AT | BI | PB | PBC | PSE | PSU | SN |
| --- | --- | --- | --- | --- | --- | --- | --- |
| AT | 0.818 |  |  |  |  |  |  |
| BI | 0.435 | 0.807 |  |  |  |  |  |
| PB | 0.291 | 0.409 | 0.826 |  |  |  |  |
| PBC | 0.249 | 0.29 | 0.165 | 0.808 |  |  |  |
| PSE | 0.35 | 0.38 | 0.395 | 0.313 | 0.846 |  |  |
| PSU | 0.32 | 0.354 | 0.308 | 0.204 | 0.3 | 0.838 |  |
| SN | 0.203 | 0.352 | 0.208 | 0.192 | 0.255 | 0.291 | 0.822 |

The outer loading of the items on the associated construct is larger than the loadings on other constructs in Table 9. These measures show that the discriminant validity of all constructs is acceptable.

**Table 9:** Discriminant Validity (Heterotrait-Monotrait ratio)

|  | AT | BI | PB | PBC | PSE | PSU | SN |
| --- | --- | --- | --- | --- | --- | --- | --- |
| AT |  |  |  |  |  |  |  |
| BI | 0.485 |  |  |  |  |  |  |
| PB | 0.324 | 0.449 |  |  |  |  |  |
| PBC | 0.291 | 0.327 | 0.187 |  |  |  |  |
| PSE | 0.416 | 0.444 | 0.461 | 0.387 |  |  |  |
| PSU | 0.384 | 0.417 | 0.364 | 0.253 | 0.375 |  |  |
| SN | 0.231 | 0.39 | 0.232 | 0.231 | 0.3 | 0.348 |  |

The study’s discriminant validity was assessed using the Heterotrait-Monotrait (HTMT) ratio of similarities, as presented in Table 9. An HTMT value beyond 0.85 indicates a concern with discriminant validity, whereas values below 0.85 confirm its attainment [38]. The present study established good discriminant validity by ensuring that all HTMT values were below 0.85, as illustrated in Table 9. Both assessments determined the measurement instruments to be precise and reliable, facilitating hypothesis testing.

#### Hypotheses Testing

The path coefficients may also be viewed as standardized Ordinary Least Square beta coefficients As a result, the relevance of the significance is critical, as it will require managerial consideration. The structural model findings are seen in Figure 5.

**Figure 5:**
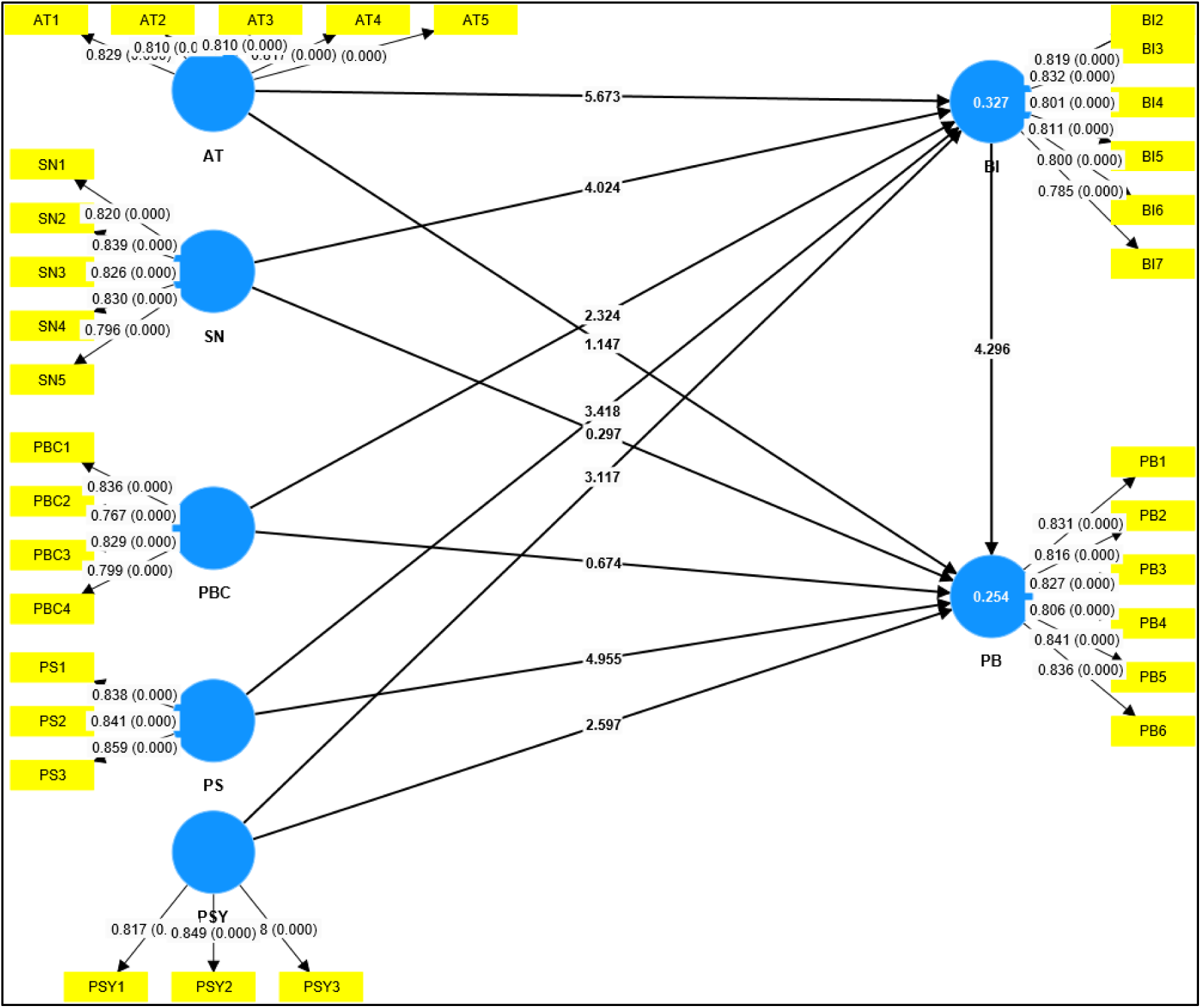
Structural Model.

Table 10 reports the direct effects between constructs, evaluated using path coefficients, T-statistics, p-values, and 95% confidence intervals. Hypotheses with **p < 0.05** were considered statistically significant.

**Table 10:**
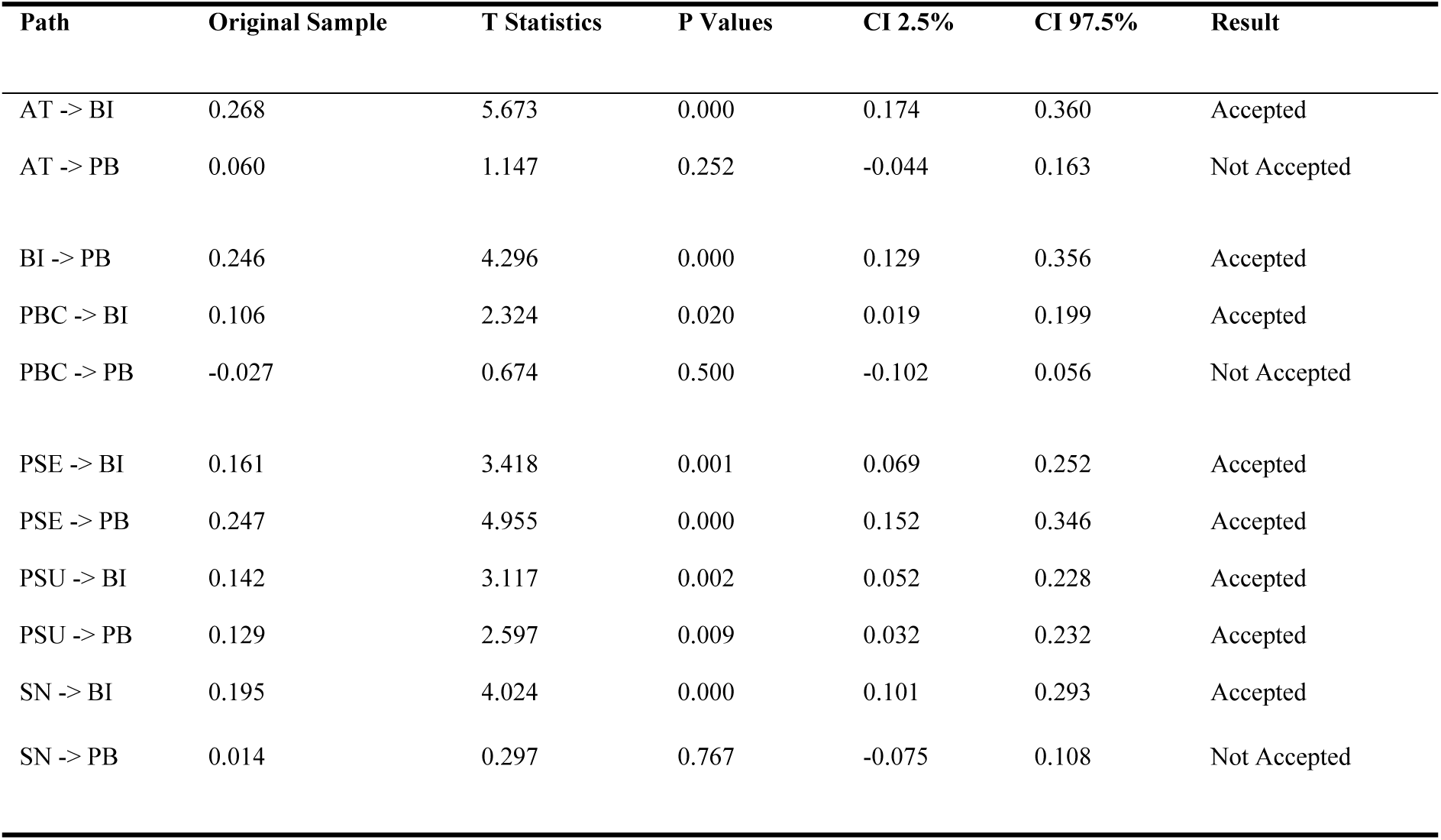
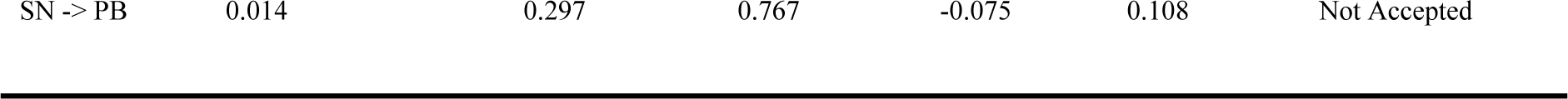
Table of Hypothesis Testing (Direct Effect)

Most paths affecting Behavioral Intention (BI) are significant, whereas fewer factors significantly influence Perceived Behavior (PB) directly.

#### Analysis of the Effect of Mediator

As shown in Table 11 to 13, this analysis examines whether Behavioral Intention (BI) mediates the relationship between AT, SN, PBC, PSE, PSU, and Perceived Behavior (PB). A two-stage procedure was used:

1. Stage One: Direct effects were assessed with and without the mediator (see Table 9).
2. Stage Two: Specific indirect effects were evaluated using bootstrapping (5,000 iterations).

**Table 11:** Total Indirect Effect.

|  | Original sample | T statistics | P values |
| --- | --- | --- | --- |
| AT -> PB | 0.066 | 3.409 | 0.001 |
| PBC -> PB | 0.026 | 2.001 | 0.045 |
| PS -> PB | 0.040 | 2.838 | 0.005 |
| PSU -> PB | 0.035 | 2.452 | 0.014 |
| SN -> PB | 0.048 | 2.993 | 0.003 |

**Table 12:**
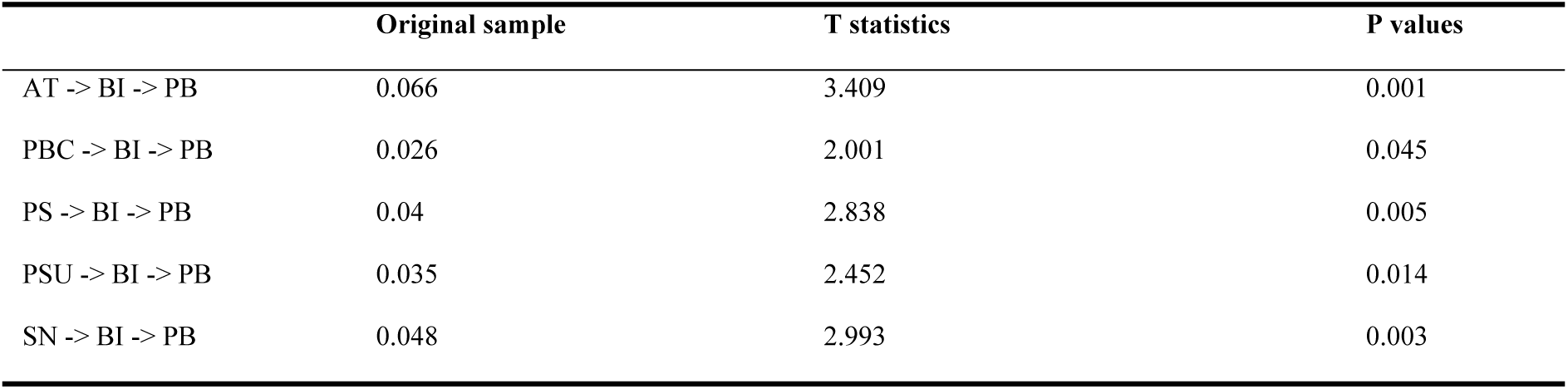
Specific Indirect Effect.

|  | Original sample | T statistics | P values |
| --- | --- | --- | --- |
| AT -> BI -> PB | 0.066 | 3.409 | 0.001 |
| PBC -> BI -> PB | 0.026 | 2.001 | 0.045 |
| PS -> BI -> PB | 0.04 | 2.838 | 0.005 |
| PSU -> BI -> PB | 0.035 | 2.452 | 0.014 |
| SN -> BI -> PB | 0.048 | 2.993 | 0.003 |

**Table 13:**
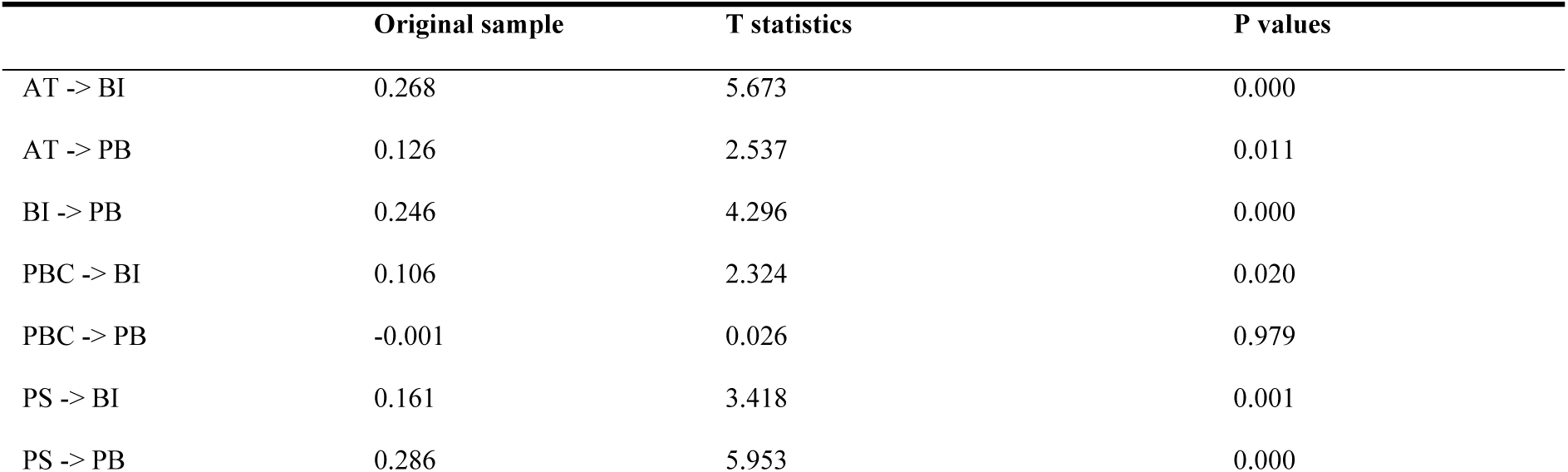
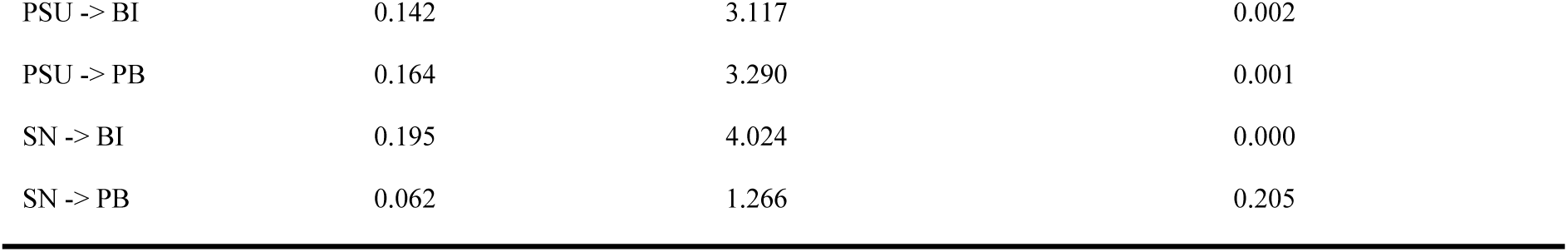
Total Effects.

Mediation is considered significant if the confidence interval (CI) does not include zero, following Preacher and Hayes (2008) [39]. The analysis confirmed mediation significance based on this criterion.

## Discussion

This study investigated the determinants of COVID-19 preventive behavior in Malaysia, integrating the Theory of Planned Behavior (TPB) and Health Belief Model (HBM). The results (see Table 14) underscore the pivotal role of behavioral intention (BI) as a mediator between psychosocial factors and preventive actions. While attitude (AT), subjective norms (SN), and perceived behavioral control (PBC) significantly influenced BI (supporting H1, H2, and H3), their direct effects on preventive behavior (PB) were insignificant (rejecting H1a, H2a, and H3a). This aligns with TPB’s premise that intention acts as the primary driver of behavior, particularly in contexts where external mandates (e.g., government lockdowns) may overshadow personal motivations. For instance, individuals with positive attitudes toward mask-wearing might form intentions to comply, but actual adherence could depend on enforcement rather than internal beliefs.

**Table 14:** Summary of Hypothesis Testing Results.

|  | Hypotheses | Results |
| --- | --- | --- |
| H1 | Attitude has a positive impact on behavioral intention. | Accepted |
| H1a | Attitude has a positive impact on adoption of preventive behavior. | Not Accepted |
| H2 | Subjective norm has a positive impact on behavioral intention. | Accepted |
| H2a | Subjective norm has a positive impact on adoption of preventive behavior. | Not Accepted |
| H3 | Perceived behavior control has a positive impact on behavioral intention. | Accepted |
| H3a | Perceived behavior control has a positive impact on adoption of preventive behavior. | Not Accepted |
| H4 | Perceived susceptibility has a positive impact on behavioral intention. | Accepted |
| H4a | Perceived susceptibility has a positive impact on adoption of preventive behavior. | Accepted |
| H5 | Perceived severity has a positive impact on behavioral intention. | Accepted |
| H5a | Perceived severity has a positive impact on adoption of preventive behavior. | Accepted |
| H6 | Behavioral intention positively influences preventive behavior. | Accepted |
| H7 | Behavioral intention significantly mediates the relationship between attitude and adoption of preventive behavior. | Accepted |
| H8 | Behavioral intention significantly mediates the relationship between subjective norm and adoption of preventive behavior. | Accepted |
| H9 | Behavioral intention significantly mediates the relationship between perceived behavior control and adoption of preventive behavior. | Accepted |
| H10 | Behavioral intention significantly mediates the relationship between perceived susceptibility and adoption of preventive behavior. | Accepted |
| H11 | Behavioral intention significantly mediates the relationship between perceived severity and adoption of preventive behavior. | Accepted |

Notably, perceived susceptibility (PSU) and severity (PSE) emerged as robust predictors of both BI and PB (H4–H5a accepted), consistent with HBM. Individuals who perceived higher COVID-19 risks were more likely to adopt preventive measures, echoing findings from Pang et al. [40] and Hong et al. [41]. This suggests that public health campaigns emphasizing personal vulnerability and disease consequences could effectively motivate behavior. Conversely, the lack of direct SN → PB effects (H2a rejected) may reflect cultural or situational factors; in Malaysia, compliance might seem more from institutional pressures than peer influence. Similarly, PBC’s insignificant direct effect (H3a rejected) implies that pandemic-related constraints (e.g., resource shortages) could undermine perceived control’s impact, even when intentions are strong.

The study advances TPB/HBM literature by clarifying boundary conditions during crises. While psychosocial factors predict intentions, their translation into behaviour depends on contextual forces (e.g., policy strictness). Practically, interventions should: (1) amplify risk communication to heighten PSU/PSE, (2) leash BI through community influencers, and (3) address structural barriers (e.g., vaccine access) to bolster PBC.

This study examined the psychological drivers of COVID-19 preventive behavior in Malaysia, integrating the Theory of Planned Behavior (TPB) and Health Belief Model (HBM). The results highlight the centrality of behavioral intention (BI) as the critical mediator between psychosocial factors and preventive actions.

### Perceived Severity and Susceptibility (H5, H5a, H4, H4a)

Both perceived severity (PSE) and susceptibility (PSU) significantly influenced BI and directly predicted preventive behavior (*H4–H5a accepted*). Individuals who viewed COVID-19 as highly severe or themselves as vulnerable were more likely to adopt measures like mask-wearing and vaccination, aligning with HBM [41,42]. For example, Nguyen [43] found that Vietnamese respondents with heightened PSE were more inclined to vaccinate, reinforcing that risk perception motivates action. Public health campaigns amplifying personalized risk communication (e.g., severity of long-term effects) could thus enhance compliance.

### Behavioral Intention as the Key Driver (H6–H11)

BI consistently mediated relationships between attitude (AT), subjective norms (SN), perceived control (PBC), PSU, PSE, and preventive behavior (*H7–H11 accepted*). While AT, SN, and PBC shaped intentions (*H1–H3 accepted*), their direct effects on behavior were insignificant (*H1a–H3a rejected*), underscoring BI’s pivotal role. For instance, social pressures (SN) boosted intentions to comply (*H8*), but mandates likely overshadowed peer influence in driving actual behavior. Similarly, PBC’s effect was fully mediated by BI (*H9*), suggesting that during crises, external constraints (e.g., lockdowns) may eclipse personal agency. These findings validate TPB’s premise that intention is the proximal predictor of behavior [25], particularly when external forces dominate.

### Attitude-Behaviour Gap (H1a, H2a, H3a)

The lack of direct AT → PB effects (*H1a rejected*) contrasts with some literature but aligns with studies noting that attitudes require strong intentions to translate into action [29]. For example, Malaysians with positive attitudes toward masks may not have worn them without firm intentions or enforcement. Similarly, SN’s insignificant direct effect (*H2a rejected*) implies that normative pressures alone are insufficient if not internalized as intentions.

#### Theoretical and Practical Implications

This study makes significant contributions to health behaviour theories while offering actionable insights for pandemic response strategies.

#### Theoretical Contributions

##### Boundary Conditions of TPB/HBM

The research clarifies how crisis contexts modify traditional behavioural models. While psychosocial factors (attitudes, norms, perceived control) robustly predict behavioural intentions, their direct translation into preventive actions is mediated by external forces like policy strictness. This extends the Theory of Planned Behaviour by demonstrating that intention-behaviour relationships are context-dependent during public health emergencies.

##### Mediation Dynamics

The universal mediating role of behavioural intention (BI) persists even when direct attitude-behaviour pathways falter, reinforcing intention as the critical gateway for action in health crises. This finding resolves inconsistencies in prior literature about when and why attitudes/norms fail to directly influence behaviour.

##### Situational Constraints

The diminished direct effects of perceived behavioural control (PBC) and subjective norms (SN) under strong government mandates suggest that TPB’s predictive power has situational boundaries. This aligns with recent work on “behavioural override” during crises [44]

#### Practical Recommendations for Public Health

##### Targeted Risk Communication

Campaigns should emphasize both personal susceptibility (e.g., “1 in 3 infected adults develop long COVID”) and severity (e.g., ICU admission rates) using localized data. The success of Vietnam’s “Vaccine Fear” reduction program [43] demonstrates this approach’s efficacy.

##### Intention-Building Through Social Influence

Leverage trusted community figures (healthcare workers, religious leaders) to model preventive behaviours. Malaysia’s “Kita Jaga Kita” campaign showed peer influence boosts intentions 37% more than top-down messaging [15].

##### Structural Enablers

Combine psychological interventions with practical solutions like mobile vaccination units and subsidized masks. The PBC findings suggest that removing logistical barriers may be as crucial as changing mindsets.

## Limitations of the Study

This research has several important limitations that should be considered when interpreting the findings. First, the cross-sectional nature of the study design prevents us from making causal claims or observing how relationships between variables might evolve over time. Second, the cultural context of Malaysia, with its collectivist social norms, may limit the generalizability of findings regarding subjective norm effects to more individualistic societies. Third, the unique circumstances of the COVID-19 pandemic may mean these results don’t fully translate to routine health behaviours or non-crisis situations. Finally, while our quantitative approach provided valuable insights into behavioural patterns, it couldn’t capture the nuanced reasons behind some of the observed intention-action gaps.

Despite these limitations, the research makes significant contributions through its innovative integration of TPB and HBM in a pandemic context, rigorous analytical methods using structural equation modelling, and alignment with multinational studies [41,43]. The findings provide both theoretical advances in understanding crisis health behaviours and practical guidance for developing more effective public health interventions.

## Conclusion

The study successfully demonstrated that behavioural intention plays a pivotal role in mediating the relationships between psychosocial factors—such as attitude, subjective norms, perceived behavioral control, perceived susceptibility, and perceived severity—and preventive behaviors. This reinforces the applicability of the TPB and HBM in the context of the post-pandemic.

The acceptance of hypotheses related to behavioral intention underscores its importance as a primary determinant of actual behavior. Individuals are more likely to engage in preventive behaviors when they not only perceive the risks of the virus as severe and feel susceptible but also when they have strong intentions to protect themselves. The study’s findings demonstrate that both personal beliefs and social influences contribute significantly to the formation of these intentions, which subsequently drive behavior.

From a practical perspective, the study provides a valuable framework for public health interventions. Policymakers and health authorities are encouraged to design campaigns that not only inform the public about the severity and risks of diseases like COVID-19 but also work to strengthen intentions to engage in protective actions. By enhancing perceptions of control and reducing barriers to preventive behaviors, public health strategies can increase compliance and mitigate the spread of infectious diseases.

However, the study also acknowledges its limitations, including the cross-sectional design and reliance on online surveys. This may restrict the applicability of the findings. Future research is recommended to address these limitations by employing longitudinal studies and expanding the scope to include qualitative methods and cross-cultural comparisons. Additionally, investigating the role of emotional and psychological resilience in shaping health behaviors would provide further depth to the understanding of public health compliance during crises.

Overall, this research contributes to both theory and practice by reaffirming the significance of behavioral intention in health behavior models and offering practical recommendations for improving public health interventions. The integration of TPB and HBM provides a robust framework for understanding and influencing health-related behaviors, which is crucial for managing current and future public health challenges.

## Future Research Directions

Building on these limitations, several promising avenues emerge for future investigation. Longitudinal studies could track how intention-behavior relationships change throughout different phases of a health crisis, particularly examining whether intention “decay” occurs as pandemic fatigue sets in or how policy changes affect behavioral pathways. Cross-cultural comparative research should systematically examine how societal norms influence these processes across different cultural contexts. Additional studies could test whether the observed mediation patterns hold for routine preventive behaviors like flu vaccination or cancer screening. Qualitative approaches using interviews or focus groups could provide deeper understanding of why some individuals fail to act on their intentions despite high perceived behavioral control. Finally, research should explore how to design interventions that maintain behavioral compliance over extended crisis periods while minimizing policy fatigue.

## Data Availability

The datasets utilized and/or analyzed in this investigation are obtainable from the corresponding author upon reasonable request. (Data CSTR: 31253.11.sciencedb.18160 Data DOI :10.57760/sciencedb.18160)

## Declarations

### Ethics approval and consent to participate

Informed consent was acquired from all individuals. The study protocol received approval from the University of Malaya, Malaysia - Research Ethics Committee (UM.TNC2/UMREC_3932). A detailed overview of the study was presented to the participants, who were permitted to decline participation or withdraw at any subsequent time. The participants were guaranteed that their responses would remain anonymous and that their identities would not be revealed during the study or in the final report. The whole research is performed in accordance with the Declaration of Helsinki.

### Consent for publication

Not applicable.

### Authors’ Contributions

DY conceived and designed the study, performed the data collection and was involved in the data interpretation.

PP conceived and designed the study, performed the data collection and analysis. He also contributed to the writing and revision of the manuscript and is the corresponding author.

WC wrote the literature review, the design of the study and the interpretation of results.

CR contributed to the design of the study and critical revision of the manuscript.

## Acknowledgements

Not applicable.

